# Effectiveness of a Molecular Tumor Board in Making Personalized Treatment Recommendations for Patients with Advanced Cancer

**DOI:** 10.64898/2026.10.01.26364445

**Authors:** Charles V. Rajadurai, Drew Harrigan, Josiane Lafleur, Soumaya Labidi, Francois Panet, Islam E. Elkholi, Andreas I. Papadakis, Aline Mamo, Adriana Aguilar-Mahecha, Susie Lau, Petr Kavan, Alan Spatz, Gerald Batist, Jennifer Friedmann, Parvaneh Fallah, Cristiano Ferrario, Mark Basik, April A. N. Rose

## Abstract

**BACKGROUND:** Next-generation sequencing (NGS) and the development of targeted therapies have revolutionized cancer treatment. Molecular Tumour Boards (MTB) serve as collaborative platforms to integrate individual clinical and molecular information to make recommendations for personalized treatment.

**METHOD:** This is a single-center, retrospective cohort study of patients presented at the Jewish General Hospital (JGH) MTB between July 2018 and Dec 2022. Clinical, molecular/pathologic data, MTB recommendations, and treatment details were collected. Treatment response was determined by radiological evaluation and physician clinical assessment. Fisher’s exact test and Log-rank tests were used to assess differences between groups.

**RESULTS:** A total of 328 patients were presented to the MTB, including 23 with repeat presentations, resulting in 355 case reviews. Tumour-agnostic biomarkers were detected in 18.9% of cases, most commonly high tumour mutational burden (TMB, 9.5%) and *HER2* amplification (4.3%). The most frequent actionable mutations were *PIK3CA* (19.8%) and *PTEN* (13.1%), often leading to recommendations for mTOR inhibitor therapy (26.0%). Overall, therapeutic recommendations were issued in 84.5% (300/355) of cases, and implemented in 43.3% (130/300). Patients who received MTB-recommended therapies achieved significantly higher objective response rates (ORR) (14.2% vs. 4.2%; P = 0.047), longer real-world progression-free survival (rwPFS) (5.6 vs. 3.5 months; HR 0.57, 95% CI 0.390–0.828; P = 0.003), and improved median overall survival (OS) (12.8 vs. 5.2 months; HR 0.60, 95% CI 0.452–0.793; P < 0.001) compared with those treated without MTB guidance.

**CONCLUSION:** MTB is effective in identifying biomarker-matched therapeutic recommendations for advanced malignancies where limited standard-of-care treatment option is available.

## 1. Introduction

Over the past two decades, significant advances in cancer genomics have improved our understanding of the molecular alterations underlying tumorigenesis and mechanisms of therapeutic resistance. Concurrently, the development of an expanding array of drugs targeting these “actionable” molecular alterations has transformed the therapeutic landscape, providing new opportunities for patients whose cancers progress despite standard-of-care treatments. In this context, molecular profiling can identify genomic alterations that may be matched with targeted therapies, provided these agents have demonstrated clinical efficacy and received regulatory approval within the same or across different tumor types (1, 2).

The integration of tumor molecular profiling into clinical practice has accelerated the adoption of personalized oncology, where treatment selection is increasingly guided by predictive biomarkers rather than tumor histology alone (3, 4). In many malignancies, biomarker-driven therapies have become established components of standard care. For example, patients with HER2-amplified breast cancer benefit from multiple HER2-directed therapies, including trastuzumab, pertuzumab, tucatinib, trastuzumab emtansine (T-DM1), and trastuzumab deruxtecan (T-DXd) (5–8). Similarly, trastuzumab combined with chemotherapy is a standard first-line treatment for *HER2*-amplified gastroesophageal cancers, while HER2-directed strategies continue to demonstrate activity in selected patients with *HER2* alterations across other tumor types, including bladder, colorectal, and lung cancers (9–11).

Experience with biomarker-matched therapies has increasingly demonstrated that effective targeted treatment strategies may transcend traditional tumor classifications, shifting the paradigm from tissue-of-origin-based therapy toward molecularly defined tumor agnostic therapy. This concept has been supported by regulatory approvals of several tumor-agnostic therapies. For example, trastuzumab deruxtecan, entrectinib, and BRAF/MEK inhibitors have been approved for tumors harboring *HER2* amplification, *NTRK/RET* fusions, and BRAF V600E mutations, respectively, regardless of primary tumor site (12). Similarly, pembrolizumab is approved for tumors characterized by high tumor mutational burden (TMB) or deficient mismatch repair (dMMR), defined by loss of expression of mismatch repair proteins including *MSH2, MLH1, PMS1, PMS2,* and *MSH6* (13). These advances have prompted further discussion regarding whether emerging biomarker-driven therapies, such as KRAS inhibitors with demonstrated activity across lung, pancreatic, and colorectal cancers, may ultimately warrant broader tumor-agnostic applications (14, 15).

Accordingly, comprehensive profiling for actionable molecular alterations represents an important therapeutic opportunity for patients with advanced cancers, particularly following failure of standard treatment options (4). This evolution reflects a fundamental paradigm shift in oncology from a one-size-fits-all approach based on tumor type toward a precision medicine model guided by molecular vulnerabilities (1, 2). Despite these advances, the efficacy of some biomarker-matched therapy varies substantially across tumour types. For example, HER2-targeted therapies yield high response rates in HER2- amplified breast cancer, but only modest responses in HER2-amplified upper gastrointestinal cancers when combined with chemotherapy (7, 9). These complexities underscore the challenge of selecting effective molecular-based therapies and highlight the need for robust frameworks to interpret genomic data and translate them into clinically meaningful treatments.

To facilitate the interpretation of complex genomic data and the integration of molecular findings with clinical context, MTBs have emerged as multidisciplinary platforms to support personalized therapeutic decision-making (3, 16, 17). The JGH MTB was the first MTB in Quebec, Canada and was established in Montreal in July 2018. Since 2018, the JGH MTB has reviewed numerous advanced cancer cases and made therapeutic recommendations beyond standard-of-care options. This study aims to characterize the impact of the of JGH MTB by examining patient demographics, treatment recommendations, clinical implementation, and patient outcomes following MTB-guided interventions.

## 2. Material and Methods

### 2.1 Study design

We conducted a retrospective cohort study of cases presented at the JGH MTB from July 2018 to December 2022. The approval for this study was obtained from the Research Ethics Board of CIUSSS de Centre-Ouest-de-l’île-de-Montréal on May 13, 2024 (Project Approval Number 2023-3621).

### 2.2. Patient characteristics

Key inclusion criteria were: patients aged 18 years and older with various types of advanced cancer who were presented at the JGH MTB between July 2018 and December 2022. We included patients from the inception of the JGH MTB (July 2018) but excluded those presented after December 2022. All patient data were censored as of May 30, 2023.

### 2.3. Data collection

Upon receiving approval from the Direction des Services Professionnels (DSP) du CIUSSS de Centre-Ouest-de-l’île-de-Montréal, using the local electronic medical records, we established a comprehensive database. This database was constructed through a systematic probabilistic sampling methodology, encompassing all patients who fulfilled our predetermined inclusion criteria.

### 2.4. Outcome Measures

The primary endpoint was Objective response rate (ORR), real-world progression free survival (rwPFS) and overall survival (OS) to evaluate the efficacy of MTB recommended therapy in patients with advanced cancer. Radiological evidence of progression had to be noted on two consecutive scans (MRI, CT-scan or PET-scan) to rule out pseudo- progression on immunotherapy. For patients whose disease progressed during MTB- recommended therapy, rwPFS was defined as the time from initiation of the treatment to disease progression. Disease Progression was defined as either the first radiographic evidence of progression accompanied by clinical determination of progression by the treating oncologist, or the second radiographic assessment confirming progression. Disease progression was determined radiologically in accordance with the iRECIST criteria and/or the clinical judgment of the radiologist and oncologist. If there was a partial response (PR) or complete response (CR) to treatment or a stable disease (SD), rwPFS was censored at the time of the last scan. OS was defined as the time between MTB recommendation and patient death if the patient was deceased, and if the patient was still alive, censored at the time of the last contact date in medical record at the time of data collection.

The secondary endpoints were to assess specific patient, disease and treatment characteristics. For the patient characteristics, demographics, clinical and pathologic data were collected. The performance status at the initiation of treatment was graded from 0 to 4 in accordance to the Eastern Cooperative Oncology Group (ECOG) noted by the oncologist. The type of MTB recommended systemic therapy as well as the number of cycles received and the line of treatment was collected.

### 2.5 Statistical Analysis

All statistical analyses were done with StataBE version 18.0.130 and Excel 2016. A p-value <0.05 was considered statistically significant for all analyses. Categorical variables and continuous variables separated into categories were presented as frequencies. Log-rank tests were used to assess differences in OS and rwPFS between groups, and median rwPFS and OS were presented along with 95% confidence intervals and Kaplan-Meier plots. Multivariate Cox regression was used to calculate hazard ratio (HR) along with confidence intervals for each chosen factors believed to predict rwPFS and/or OS. ECOG (≤1 or >1), number of lines of therapy (≤3 or > 3) and age (≥ 70 or < 70) were considered as categorical variables with the reference value. Fisher’s exact test was used to evaluate ORR between groups.

## 3. Results

### 3.1 Molecular Tumour Board Design

The JGH MTB comprises medical and surgical oncologists, pathologists, scientists, clinical biology specialists, clinical and research trainees, and clinical trial coordinators. Treating oncologists submit cases of advanced cancer for discussion. Pathology reports, biomarker profiles, and molecular diagnostics are reviewed, and the MTB provides a consensus treatment recommendation. Access to the proposed therapy was then pursued through standard healthcare coverage, special access programs (e.g., compassionate use), or clinical trial enrollment. If the patient could not receive the recommended treatment— due to drug inaccessibility or physician/patient choice—the case could be re-presented at a future MTB (**Figure 1**).

**Figure 1.**
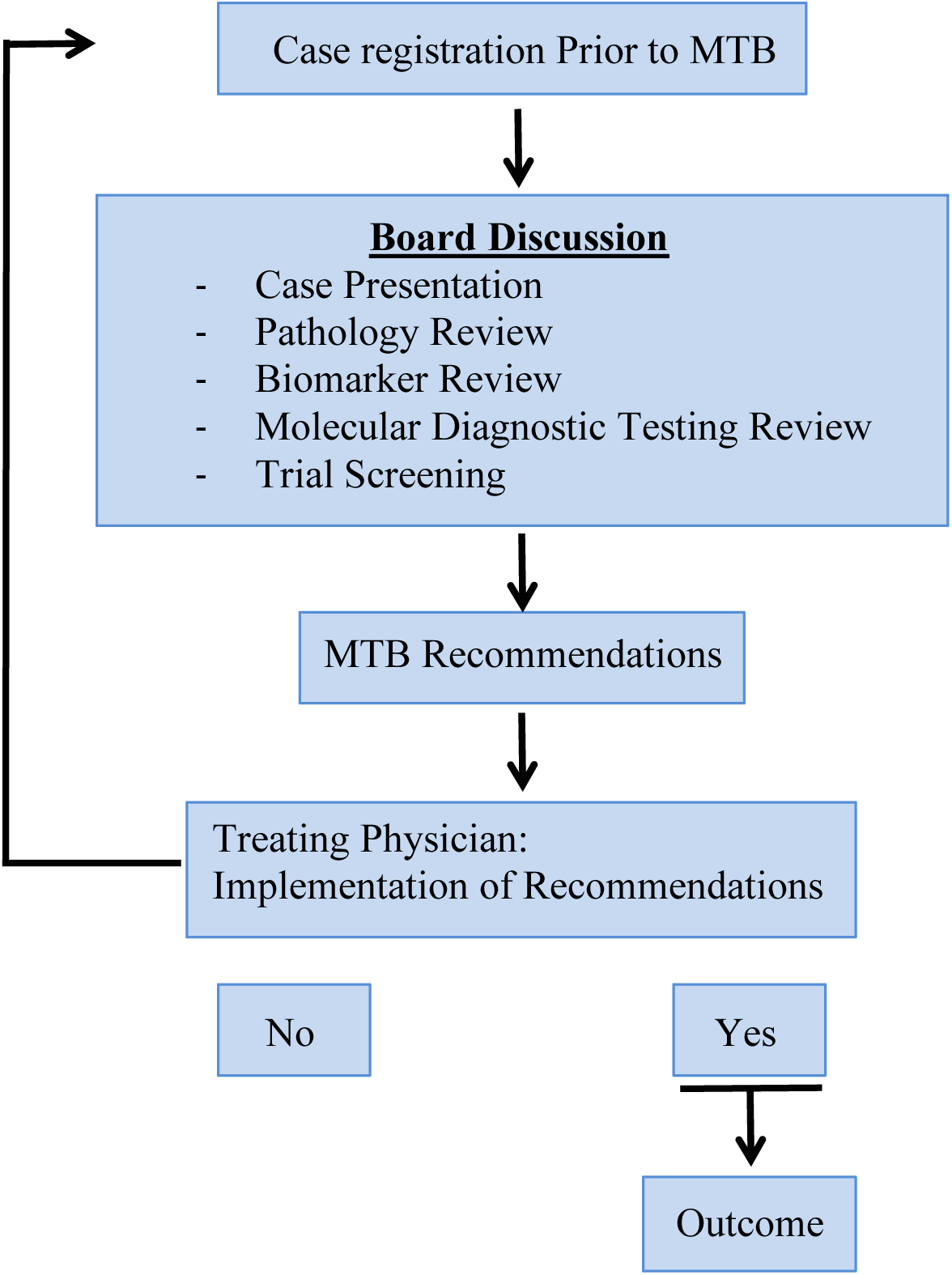

### 3.2 Patient Characteristics

A total of 328 advanced cancer patients were presented to the JGH MTB between July 2018 and December 2022. The majority was female (58.8%; 193/328), with a median age of 62 years (range: 23–101) (**Table 1)**. Genitourinary cancers were the most common (25.0%; 82/328), followed by breast (21.3%; 70/328), gastrointestinal (20.4%; 67/328), and gynecological cancers (18.9%; 62/328) (**Table 1)**. Nearly all cases (98.5%; 323/328) were stage IV at the time of MTB presentation (**Table 1)**. Most patients had an ECOG performance status of 1 (78.0%; 256/328), with 15.5% (51/328) at ECOG 2, 5.2% (17/328) at ECOG 0, and 1.2% (4/328) at ECOG 3 (**Table 1**).

**Table 1:** MTB Patient Characteristics

| Number of Patients | 328 |
| --- | --- |
| Clinical Characteristic | N (%) |
| Sex |  |
| - F | 193 (58.8) |
| - M | 135 (41.2) |
| Age, Median (Range) | 62 (23-101) |
| Tumor Site |  |
| - GU | 82 (25.0) |
| - Breast | 70 (21.3) |
| - GI | 67 (20.4) |
| - Gynecological | 62 (18.9) |
| - Lung | 17 (5.2) |
| - Other | 30 (9.1) |
| Stage |  |
| - III | 5 (1.5) |
| - IV | 323(98.5) |
| ECOG |  |
| - 0 | 17 (5.2) |
| - 1 | 256 (78.0) |
| - 2 | 51 (15.5) |
| - 3 | 4 (1.2) |

### 3.3 Molecular Diagnostics and Biomarker Review

Among patients presented at the MTB, 78.7% (258/328) underwent next-generation sequencing (NGS) via tissue biopsy, 18.6% (61/328) via circulating tumor DNA (ctDNA) from liquid biopsy, and 2.7% (9/328) via both methods (**Supplemental Figure 1)**. The Foundation One panel was the most commonly used NGS platform for both tissue (97.8%; 261/267) and liquid biopsies (91.4%; 64/70). For the remaining liquid biopsies, Canexia (5.7%; 4/70) and Guardant360 (2.9%; 2/70) were the only other panels used (**Supplemental Figure 1**).

Of the patients presented at the JGH MTB, 18.9% (62/328) had at least one molecular alteration with an FDA-approved indication for a tumour-agnostic, biomarker-matched therapy (**Figure 2**). Specifically, 9.5% (31/328) had high tumour mutational burden (TMB), 4.3% (14/328) had *ERBB2* amplification, 2.4% (8/328) had *BRAF* V600E mutations, 1.5% (5/328) had mismatch repair deficiency, 0.9% (3/328) had *NTRK* fusions, and 0.3% (1/328) had a *RET* fusion (**Figure 2**).

**Figure 2A).**
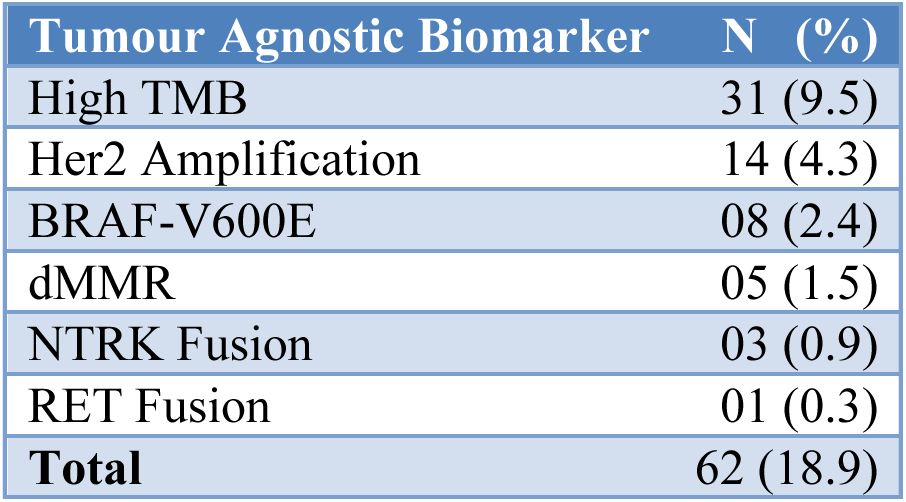

**Figure 2B).**
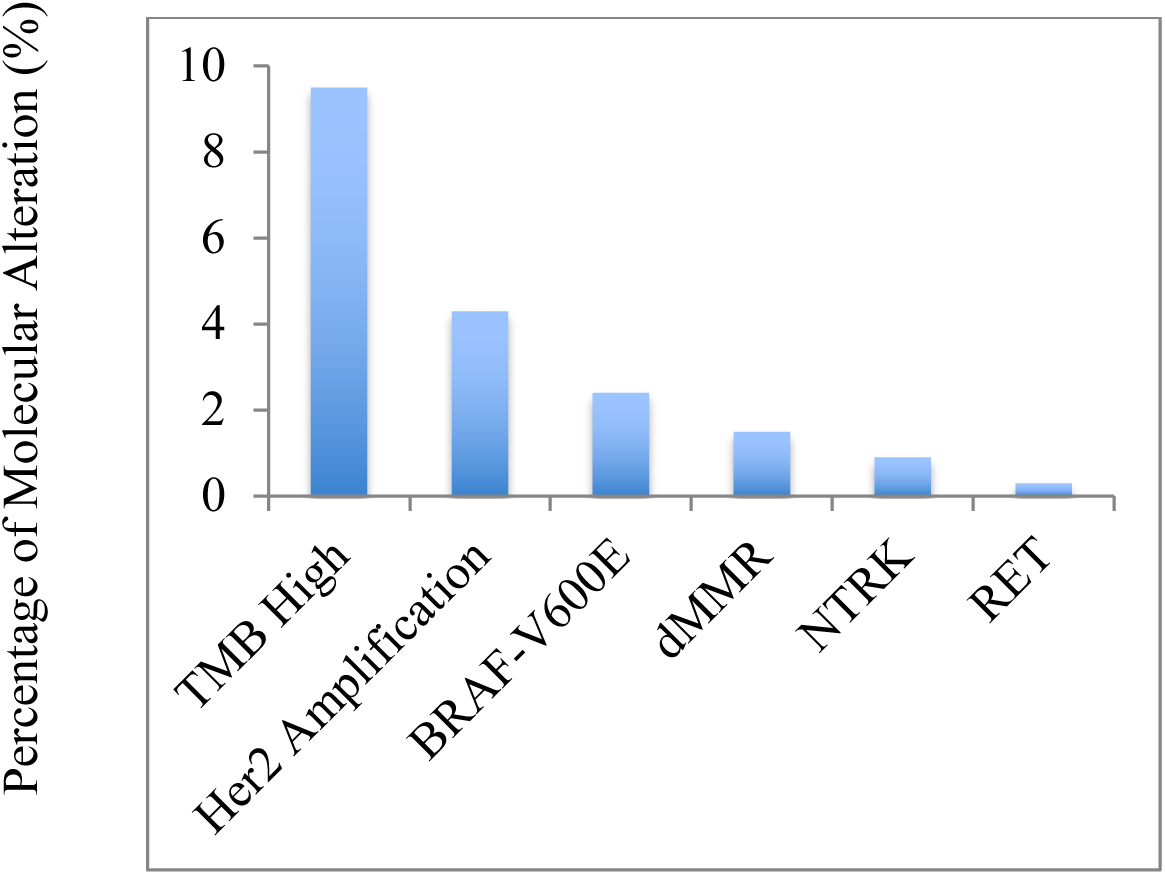

### 3.4 Molecular Alteration Review

NGS analysis identified *MYC* (11.3%, 37/328), *CCND1* (7.9%, 26/328), and *RAD21* (7.3%, 24/328) as the most frequently amplified genes (**Supplemental Figure 2A**). The most commonly deleted genes were *CDKN2A/B* (7.0%, 23/328), *PTEN* (2.7%, 9/328), and *MTAP* (2.4%, 8/328) (**Supplemental Figure 2A**). TP53 was the most frequently mutated gene, observed in 53.7% of cases (176/328) (**Supplemental Figure 2B**). Among actionable mutations, the most prevalent were *PIK3CA* (18.3%, 60/328), *KRAS* (13.4%, 44/328), and *PTEN* (12.8%, 42/328) (**Supplemental Figure 2B**). Frequent molecular alterations (mutation and copy number alteration) associated with mTOR, AKT, or PIK3CA inhibitor recommendations included mutations in *PIK3CA* (18.3%; 65/328) and *PTEN* (13.1%; 43/328) (**Supplemental Figure 2C**). CDK4/6 inhibitor recommendations were most commonly based on *CDKN2A/B* (6.7%; 22/328) and *CCND1* (3.4%; 11/328) alterations (**Supplemental Figure 2C**). TKI recommendations were driven by *FGFR1* (6.1%; 20/328) and *MET/HGF* (3.0%; 10/328) alterations, while *ERBB2* (4.6%; 15/328) was the primary target for HER2-directed therapies (**Supplemental Figure 2C**). PARP inhibitors were most often recommended for alterations in *ATM* (5.2%; 16/328) and *BRCA2* (4.0%; 13/328) (**Supplemental Figure 2C**). BRAF/MEK inhibitor recommendations are most frequently based on *BRAF* alterations (2.7%; 8/328) (**Supplemental Figure 2C**).

### 3.5 Molecular Tumour Board Presentation

Of the 328 patients, 305 were presented once, 19 twice, and 4 three times, totaling 355 case presentations. Most cases (85.6%; 304/355) were presented due to disease progression on standard therapy, while 14.4% (51/355) were presented despite remaining standard treatment options (**Table 2**). At the time of presentation, 39.2% (139/355) had received ≥ 4 prior lines of therapy, while 60.8% (216/355) had received 1-3 lines of therapy (**Table 2**). At least one therapeutic recommendation was made in 84.5% of cases (300/355), with two recommendations in 50.1% (178/355) (**Table 2**). At least one recommendation was subsequently implemented in **43.3%** of cases (130/300), whereas both recommendations were implemented in only **9.0%** of cases with two recommendations (16/178) during the course of treatment (**Table 2**). Among implemented therapies, 70.5% (103/146) were accessed through clinical trials (**Table 2**). In cases where no therapy was implemented, the main reasons were: lack of access (37.1%; 63/170), poor performance status (30.6%; 52/170), patient/physician preference (17.6%; 30/170), and ongoing response to current therapy (10.6%; 18/170) (**Table 2**). Second recommendations were not implemented due to inaccessibility (51.2%; 83/162), poor performance status (48.1%; 78/162), and ongoing response (0.7%; 1/162) (**Table 2**).

**Table 2:** MTB Case Presentations

|  |  |
| --- | --- |
| **Number of Patients** | **328** |
| **MTB Presentations** | **355** |
| - Single | 305 |
| - Two | 19 |
| - Three | 4 |
|  | N (%) |
| **Reason for TB** |  |
| - Progressive disease | 304 (85.6) |
| - Treatment Option | 51 (14.4) |
| **Lines of Systemic Treatment** |  |
| - $\leq 1$ | 52 (14.6) |
| - 2 | 74 (20.8) |
| - 3 | 90 (25.4) |
| - 4 | 70 (19.7) |
| - $\geq 5$ | 69 (19.4) |
| **Therapeutic Recommendation (328 Patients)** | **355** |
| One | 122 (34.4) |
| Two | 178 (50.1) |
| None | 55 (15.5) |
| **Implementation (276 Patients)** | **300** |
| One (105 Patients) | 114 (38.0) |
| Two (11 Patients) | 16 (5.3) |
| None (156 Patients) | 166 (55.3) |
| Unknown (4 Patients) | 4 (1.3) |
| **Access to Treatment** |  |
| **First Choice** | **130** |
| - Clinical Trial | 91 (70.0) |
| - Off Clinical Trial | 39 (30.0) |
| **Second Choice** | **16** |
| - Clinical Trial | 12 (75.0%) |
| - Off Clinical Trial | 04 (25.0) |
| **Reason for Not Implementing** |  |
| **First Choice** | **170** |
| - Access to Treatment | 63 (37.1) |
| - Poor Performance Status | 52 (30.6) |
| - Physician/Patient Choice | 30 (17.6) |
| - Ongoing Response | 18 (10.6) |
| - Unknown | 7 (4.1) |
| **Second Choice** | **162** |
| - Access to Treatment | 83 (51.2) |
| - Poor Performance Status | 78 (48.1) |
| - Ongoing Response | 1 (0.7) |

### 3.6 Molecular Tumour Board Recommended Treatment Characteristics

Based on molecular diagnostics and pathology reviewed at the MTB, the most frequently recommended first-choice targeted therapies were mTOR inhibitors (26.0%, 78/300), followed by PARP inhibitors (15.3%, 46/300), tyrosine kinase inhibitors (TKIs) (15.0%, 45/300), immune checkpoint inhibitors (ICIs) (12.3%, 37/300) and CDK4/6 inhibitors (10.0%, 30/300) (**Supplemental Table 1**). Among the first-choice therapies that were implemented, the most frequently administered were mTOR inhibitors (22.3%, 29/130), followed by ICIs (20.0%, 26/130), PARP inhibitors and CDK4/6 inhibitors (each 13.8%, 18/130) and TKIs (10.8%, 14/130) (**Supplemental Table 1**).

For second-choice targeted therapies, the most frequent recommendations included mTOR inhibitors (19.1%, 34/178), TKIs (17.4%, 31/178), PARP inhibitors (14.0%, 25/178), PIK3CA/AKT inhibitors (9.6%, 17/178), and ICIs and CDK4/6 inhibitors (each 8.4%, 15/178) (**Supplemental Table 2**). Among the implemented second-choice therapies, the most commonly used were PARP inhibitors (31.3%, 5/16), followed by mTOR inhibitors and ICIs (each 18.8%, 3/16) (**Supplemental Table 2**).

### 3.7 Effectiveness of MTB Recommended Therapy

The first-choice treatment recommended by the MTB resulted in a complete response (CR) in 4.3% (5/116) and a partial response (PR) in 8.6% (10/116) of patients, yielding an overall response rate (ORR) of 12.9% (15/116) (**Table 3**). Disease stabilization was achieved in 39.7% (46/116) of patients who received the MTB-recommended first-choice treatment (Table 3). Among patients who did not receive MTB-recommended therapy but subsequently received another line of systemic treatment, the ORR was 4.2% (2/48), all PRs, with disease stabilization observed in 19.9% (31/156) (**Table 3**). The first and second choice MTB recommended treatment resulted in a combined ORR of 14.2% (18/126) compared to 4.2% (2/48) in patients who did not receive MTB-recommended therapy but received a subsequent line of systemic treatment (1-sided Fisher’s Exact P = 0.047).

**Table 3:** Clinical Outcome of Patients Treated with MTB Recommended Therapy.

| Description | N (%) |
| --- | --- |
| Outcome with 1<sup>st</sup> MTB Treatment (116 Patients; 130 Cases) |  |
| - Complete Response | 5 (4.3) |
| - Partial Response | 10 (8.6) |
| - Stable Disease | 31 (26.7) |
| - Disease Progression | 69 (59.5) |
| - Unknown | 1 (0.9) |
| Outcome with 2<sup>nd</sup> MTB Treatment (11 Patients; 16 Cases) |  |
| - Partial Response | 3 (27.3) |
| - Stable Disease | 5 (45.4) |
| - Disease Progression | 3 (27.3) |
| Subsequent Treatment (48 Patients; 55 Cases) |  |
| - Partial Response | 2 (4.2) |
| - Stable Disease | 14 (29.2) |
| - Disease Progression | 32 (66.7) |

### 3.8. Exceptional Responses

Several patients with heavily pre-treated metastatic disease experienced exceptional responses to MTB-recommended targeted therapies. An ovarian cancer patient with a *BRAF* V600E mutation, treated in the second line with the BRAF/MEK inhibitor combination, achieved a rwPFS of more than 37 months and remains alive at the time end- point of the study (**Table 4**). A patient with metastatic castration-resistant prostate cancer (mCRPC) and high tumour mutational burden (TMB), treated in the third line with the Anti-PD-L1/CTLA4 combination immune checkpoint inhibitor, achieved a prolonged rwPFS of 34 months and is also alive at end-point of the study (**Table 4**). A triple-negative breast cancer (TNBC) patient harbouring PD-L1 and PD-L2 amplifications, treated in the fourth line with the Anti-PD-L1 immune checkpoint inhibitor, achieved a CR and a rwPFS exceeding 26 months; the patient is alive at end-point of the study (**Table 4**). A gastric cancer patient with an *ATM* mutation, treated in the fifth line with the PARP inhibitor, achieved a rwPFS of more than 22 months and remains alive at end-point of the study (**Table 4**). A high grade endometrial stromal sarcoma patient with an *NTRK1* fusion, treated in the second line with a TKI, achieved a rwPFS of over 22 months and is alive at end-point of the study (**Table 4**). A bladder cancer patient with *ERBB2* amplification, treated in the third line with a HER2-targeted antibody-drug conjugate (ADC), achieved a rwPFS of more than 21 months and remains alive at end-point of the study (**Table 4**). A non-small cell lung cancer (NSCLC) patient with *CDKN2A/B* deletion, treated in the fifth line with a CDK4/6 inhibitor, achieved a rwPFS exceeding 20 months (**Table 4**). A pancreatic cancer patient with *PTEN/TSC2* mutations, treated in the fourth line with an mTOR inhibitor, achieved a rwPFS of 15 months (**Table 4**).

**Table 4:** Characteristics of Exceptional Responders to MTB Recommended Therapy

| Diagnosis | Number of Prior Lines | Biomarker | Treatment | PFS (Months) | Survival Status |
| --- | --- | --- | --- | --- | --- |
| Ovarian Cancer | 1 | BRAF-V600E | BRAF/MEK Inhibitor | > 37 | Alive |
| mCRPC | 2 | MLH1 Mutation/<br>TMB High | Anti-PD-L1/CTLA4 | > 34 | Alive |
| TNBC | 3 | PD-L1/PD-L2<br>Amplification | Anti-PD-L1 | > 26 | Alive |
| Gastric Cancer | 4 | ATM Mutation | PARP Inhibitor | > 22 | Alive |
| Endometrial Cancer | 1 | NTRK1 Fusion | Tyrosine Kinase Inhibitor | > 22 | Alive |
| Bladder Cancer | 2 | ERBB2 Amplification | Her2 Antibody-Drug Conjugate | > 21 | Alive |
| NSCLC | 4 | CDKN2A/B Deletion | CDK4/6 Inhibitor | > 20 | Dead |
| Pancreatic Cancer | 3 | PTEN/ TSC2 Mutation | mTOR Inhibitor | > 15 | Dead |

### 3.9 Overall Survival

The overall survival of patients who received the MTB recommended therapy is 12.8 months compared to 5.2 months for patients who did not receive the MTB recommended therapy (**Figure 4** **& Supplemental Figure 4**). In the multivariate Cox regression analysis for overall survival, receiving MTB-recommended treatment was independently associated with a significantly lower hazard of death (HR 0.60 [0.452-0.793]; P < 0.001) (**Table 6**). ECOG performance status ≤ 1 was also a strong independent predictor (HR 0.31 [0.218- 0.430]; P < 0.001) (**Table 6**). The progression free survival of patients who received the MTB recommended therapy is 5.6 months compared to 3.5 months for patients who did not receive MTB-recommended therapy but received a subsequent line of systemic therapy (**Figure 3** **& Supplemental Figure 3**). In the multivariate Cox regression analysis for progression free survival, receiving MTB-recommended treatment was independently associated with a significantly lower hazard of death (HR 0.57 [0. 390-0.828]; P=0.003) (**Table 5**). ECOG performance status ≤ 1 was also a strong independent predictor (HR 0.44 [0.272-0.717] (**Table 5**). Other covariates, including age, sex, stage, cancer type and number of prior lines of therapy are not significantly associated with OS or rwPFS (**Table 5** **&** **Table 6**).

**Figure 3.**
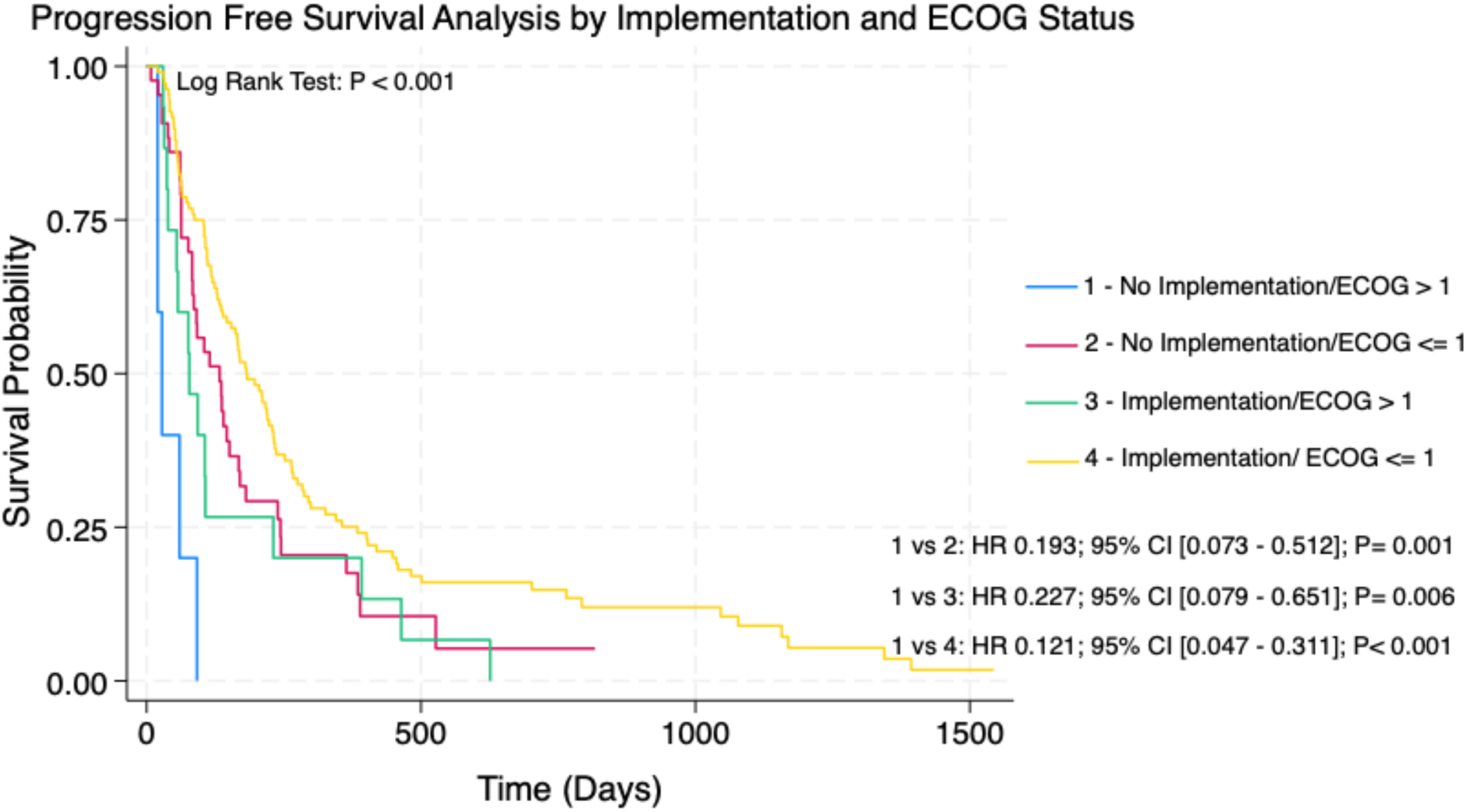

**Figure 4.**
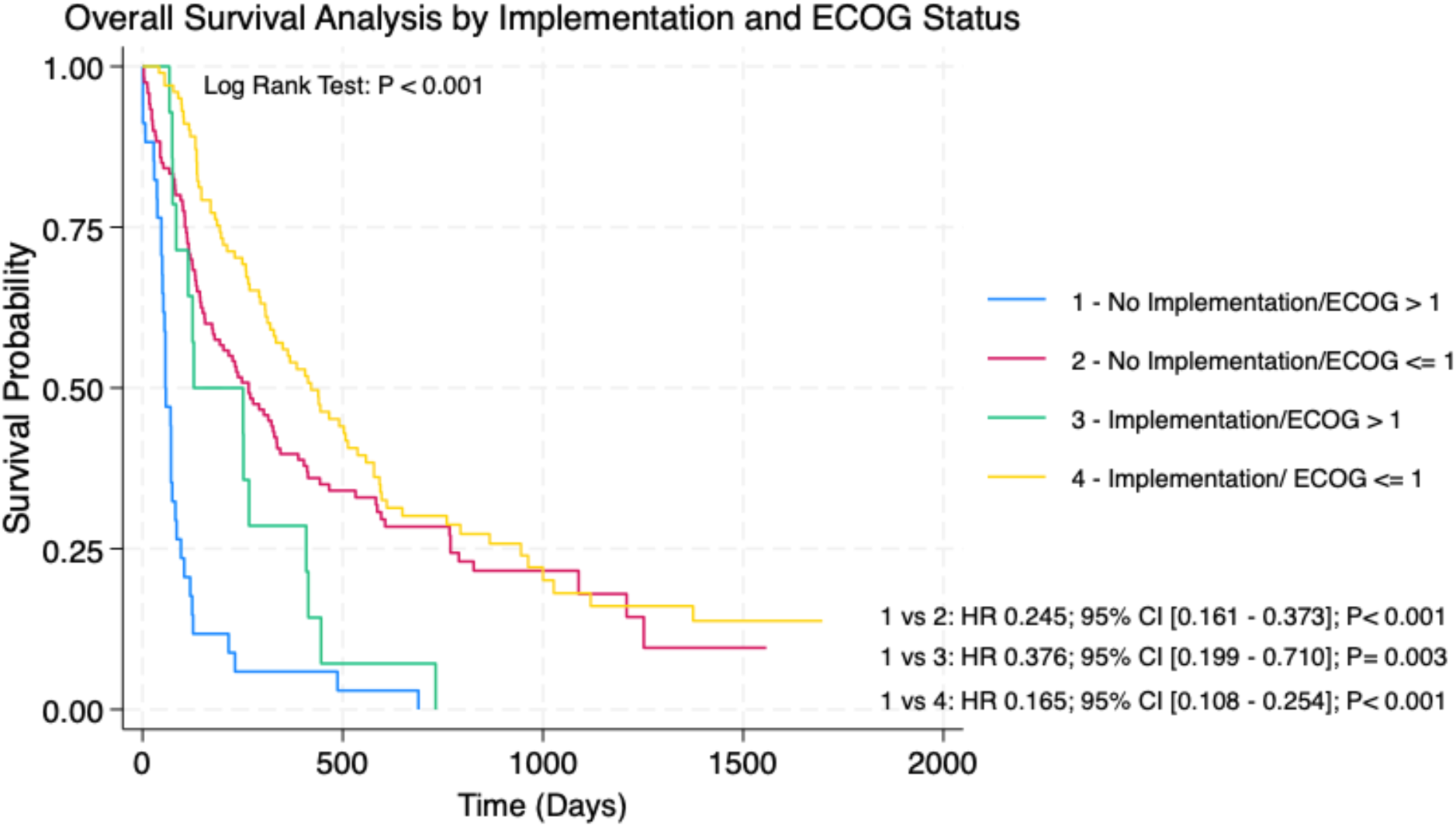

**Table 5:** Multivariate COX Regression Analysis of PFS

| Variable | Hazard Ratio | P-Value | Confidence Interval |
| --- | --- | --- | --- |
| Sexe | 1.042 | 0.788 | [0.770 - 1.411] |
| Age ($\geq 70$ vs. $< 70$) | 1.029 | 0.867 | [0.736 - 1.438] |
| Cancer (Breast & Prostate vs. Rest) | 0.810 | 0.152 | [0.607 - 1.081] |
| Stage (III vs. IV) | 3.597 | 0.206 | [0.495 - 26.15] |
| PS (ECOG $\leq 1$ vs. ECOG $>1$) | 0.306 | $< 0.001$ | [0.218 - 0.430] |
| Prior Treatment ($\leq 3$ vs. $>3$) | 0.988 | 0.936 | [0.737 - 1.324] |
| Implementation | 0.599 | $< 0.001$ | [0.452 - 0.793] |

**Table 6:** Multivariate COX Regression Analysis of OS

| Variable | Hazard Ratio | P-Value | Confidence Interval |
| --- | --- | --- | --- |
| Sexe | 0.909 | 0.595 | [0.641 - 1.291] |
| Age ($\geq 70$ vs. $< 70$) | 0.984 | 0.934 | [0.665 - 1.455] |
| Cancer (Breast & Prostate vs. Rest) | 0.848 | 0.337 | [0.606 - 1.187] |
| Stage (III vs. IV) | 0.518 | 0.376 | [0.121 - 2.223] |
| PS (ECOG $\leq 1$ vs. ECOG $>1$) | 0.441 | 0.001 | [0.272 - 0.717] |
| Prior Treatment ($\leq 3$ vs. $>3$) | 0.899 | 0.545 | [0.637 - 1.268] |
| Implementation | 0.568 | 0.003 | [0.390 - 0.828] |

## 4. Discussion

Advancements in NGS and the growing availability of biomarker-matched therapies and biomarker-defining technologies have driven the establishment of MTBs in major cancer centers worldwide(16–19). However, MTBs require significant coordination of resources, including expert personnel for molecular data interpretation and substantial funding for NGS testing and treatment access (18, 19). As a result, MTBs remain difficult to implement in resource-limited settings. To our knowledge, the JGH MTB, operational since July 2018, is the first of its kind in Quebec, Canada.

The distribution of cancer types discussed largely reflected disease prevalence (GU, breast, and GI); however, lung cancer cases were underrepresented (5.2%, 17 cases), likely because thoracic oncologists—most experienced in precision medicine—had already implemented available targeted therapies as part of standard-of-care treatment. Nearly all patients (98.8%) had an ECOG performance status of 0–2, consistent with good oncology practice (**Table 1**). Approximately 65% of patients were presented at the MTB after receiving two lines of therapy, often after exhausting or while receiving last line of standard-of-care treatment. This aligns with the fact that the predominant cancer types discussed at JGH MTB (GU, breast, GI, and gynecologic) often lack actionable targets outside of ERBB2 (in breast and gastric cancer), PIK3CA (in breast cancer) and BRAF V600E (in colorectal cancer). The MTB recommendation rate (∼85%) and implementation rate (∼43%) are notably higher than reported in the literature (**Table 2**) (20–22). Since most recommended therapies are not covered by public healthcare, around 70–75% of both first and second-line recommendations were accessed through clinical trials. Inaccessibility accounted for ∼37% of unimplemented first-line and ∼51% of second-line recommendations, while patient deterioration accounted for ∼30% and ∼48%, respectively (**Table 2**). Earlier case presentation may prevent treatment delays and functional decline, as MTB review and therapy access typically require time.

Consistent with the study period, most NGS tests were performed on solid tumour samples using the commercially available Foundation One panel. Liquid biopsies (<20%) were primarily conducted after 2021, coinciding with advancements in ctDNA technology (**Supplemental Figure 1**). A notable proportion (∼19%) of tumour-agnostic, biomarker-matched therapies were identified in a heavily pre-treated population lacking standard care options (**Figure 2**). The majority (∼14%) involved high TMB or *HER2* amplification—biomarkers linked to therapies like Pembrolizumab and T-DXd, which are widely used and well-tolerated in clinical practice (Figure 2).

Not surprisingly, *TP53* was the most frequently mutated gene, while *MYC* was the most commonly amplified (Supplementary Figure 2). Both remain largely undruggable to date, with the exception of the *TP53*-Y220C mutant, which is targeted by rezatopt and is currently being evaluated in the phase II trial PYNNACLE (23). Alterations in *PIK3CA*, *PTEN*, and *AKT* led to the highest number of targeted therapy recommendations, primarily involving mTOR and PIK3CA inhibitors (**Supplemental Figure 2**). The molecular alteration profile characterized in this study is concordant with those reported by other MTB studies (22, 24). Only three patients received AKT inhibitors, mostly through clinical trials. Among kinases, *ERBB2*, *FGFR1*, and *MET* were frequently altered, with ERBB2- targeted therapies having the highest implementation rate, likely due to greater accessibility (**Supplemental Figure 2** & **Supplemental Table 1**). Immunotherapy was frequently implemented in cases with high TMB, PD-L1/PD-L2 amplification or mismatch repair gene alterations (*PMS2*, *MLH1*, *MSH2*, *MSH6*). PARP inhibitors were recommended for *ATM*, *BRCA2*, *CHEK2*, and *PALB2* alterations, while CDK4/6 inhibitor recommendations were mostly based on *CDKN2A/B* loss (**Supplemental Figure 2**).

The implementation rate of first-line MTB recommendations (43%) was significantly higher than for second-line recommendations (9%), underscoring the need to prioritize optimal treatment early (**Table 2).** Barriers to implementation likely include declining performance status and limited treatment access. Remarkable and durable responses were observed in several cases treated with non-standard, biomarker-matched therapies (**Table 4**). A metastatic TNBC patient with PD-L1/PD-L2 gene amplification and a patient with mCRPC harbouring an *MLH1* mutation both showed excellent responses to immunotherapy (**Table 4).** Notably, the TNBC patient had a PD-L1 CPS of 5%, below the threshold for standard-of-care immunotherapy(25), highlighting the limits of immunohistochemical measurements. Heavily pre-treated cases of gastric, NSCLC, and pancreatic cancers also responded well to Olaparib (*ATM* alteration), Palbociclib (*CDKN2A/B* loss), and Temsirolimus (*PTEN/TSC2* mutations), respectively, despite these therapies not being standard care (**Table 4**). These cases illustrate that “Lazarus” responses are possible with biomarker-guided treatment, even in refractory settings.

Although the objective response rate (ORR) among patients receiving MTB- recommended therapies was modest (14.2%) compared to 4.2% in those treated with their oncologist’s choice (**Table 3**), this difference may be clinically meaningful given the heavily pre-treated nature of the cohort. Patients treated with MTB-recommended therapies achieved a median OS of 12.8 months (HR 0.60; *P* < 0.001) and a median rwPFS of 5.6 months (HR 0.57; *P* = 0.003) (Figures 4 and 5), corresponding to a 40% reduction in the risk of death or disease progression—again clinically significant in this challenging setting. Good performance status emerged as an independent predictor of improved OS and rwPFS in this group, underscoring the importance of careful patient selection. In contrast, neither cancer type (including those with more standard-of-care options) nor the number of prior treatment lines had a statistically significant impact on OS or rwPFS among patients receiving MTB-recommended therapies.

## 5. Limitations

This study is limited by its retrospective, single-center design, which is subject to selection bias, incomplete data capture, and unmeasured confounding. As treatment recommendations and subsequent management were based on routine clinical practice rather than a standardized protocol, factors influencing implementation may not have been fully captured. Furthermore, the findings reflect the experience of a tertiary academic center and may not be generalizable to other practice settings. The modest sample size also limits the precision of subgroup analyses. Accordingly, these results should be interpreted as exploratory and hypothesis-generating, warranting validation in larger, prospective multicenter studies (26).

## 6. Conclusion

The organization of a Molecular Tumour Board (MTB) requires a coordinated effort among healthcare professionals, scientists, and research teams, along with significant financial resources. MTBs are particularly beneficial for fit patients with advanced malignancies lacking standard treatment options, potentially providing additional treatment options (22, 24). Even in heavily pre-treated cases, biomarker-matched therapies can yield durable responses. Early referral to MTB may prevent delays that lead to performance status decline and missed therapeutic opportunities. As NGS technology advances and costs decrease, MTBs are becoming increasingly viable across cancer centers. However, improved strategies are needed to ensure timely access to the repertoire of emerging targeted therapies.

## Data Availability

All data produced in the present work are contained in the manuscript

## 7. Acknowledgements

IEE is supported by a Canadian Cancer Society Postdoctoral Award (#708442). A.A.N.R. acknowledges salary support from a Fonds de Recherche du Québec—Santé (FRQS) Clinical Research Scholar Award.

## Ethical approval

Project 2023-3621 is approved by CIUSSS West-Central Montreal. Final authorization to conduct research was obtained on 13 May 2024.

## Author Contributions

C.V.R. designed the research protocol, performed data collection and analysis, and drafted the manuscript. A.A.N.R. conceived the study, supervised the project, and reviewed and revised the manuscript. M.B. and C.F., co-founders of the JGH Molecular Tumour Board (MTB), regularly contributed cases to the MTB and participated in the manuscript review. D.H. contributed to data collection. J.L. coordinated the MTB case presentations and authored the MTB reports. S.L., P.K., J.F., G.B., P.F., F.P., and S.L. frequently presented cases to the MTB and contributed to the manuscript review. A.A., A.P., A.S., I.E., and A.M. participated in the interpretation of the data at MTB.

## References

1. Chakravarty D, Johnson A, Sklar J, Lindeman NI, Moore K, Ganesan S, et al. Somatic Genomic Testing in Patients With Metastatic or Advanced Cancer: ASCO Provisional Clinical Opinion. J Clin Oncol. 2022;40(11):1231–58.

2. Jamalinia M, Weiskirchen R. Advances in personalized medicine: translating genomic insights into targeted therapies for cancer treatment. Ann Transl Med. 2025;13(2):18.

3. Hoefflin R, Geissler AL, Fritsch R, Claus R, Wehrle J, Metzger P, et al. Personalized Clinical Decision Making Through Implementation of a Molecular Tumor Board: A German Single-Center Experience. JCO Precis Oncol. 2018;2.

4. Denisenko TV, Ivanova AE, Koval A, Silachev DN, Jia L, Sukhikh GT, et al. Signalomics for molecular tumor boards and precision oncology of breast and gynecological cancers. Mol Syst Biol. 2025.

5. Cortes J, Kim SB, Chung WP, Im SA, Park YH, Hegg R, et al. Trastuzumab Deruxtecan versus Trastuzumab Emtansine for Breast Cancer. N Engl J Med. 2022;386(12):1143–54.

6. Murthy RK, Loi S, Okines A, Paplomata E, Hamilton E, Hurvitz SA, et al. Tucatinib, Trastuzumab, and Capecitabine for HER2-Positive Metastatic Breast Cancer. N Engl J Med. 2020;382(7):597–609.

7. Swain SM, Baselga J, Kim SB, Ro J, Semiglazov V, Campone M, et al. Pertuzumab, trastuzumab, and docetaxel in HER2-positive metastatic breast cancer. N Engl J Med. 2015;372(8):724–34.

8. von Minckwitz G, Huang CS, Mano MS, Loibl S, Mamounas EP, Untch M, et al. Trastuzumab Emtansine for Residual Invasive HER2-Positive Breast Cancer. N Engl J Med. 2019;380(7):617–28.

9. Bang YJ, Van Cutsem E, Feyereislova A, Chung HC, Shen L, Sawaki A, et al. Trastuzumab in combination with chemotherapy versus chemotherapy alone for treatment of HER2-positive advanced gastric or gastro-oesophageal junction cancer (ToGA): a phase 3, open-label, randomised controlled trial. Lancet. 2010;376(9742):687–97.

10. Killock D. HER2-directed therapy improves urothelial cancer outcomes. Nat Rev Clin Oncol. 2025;22(12):898.

11. Li BT, Smit EF, Goto Y, Nakagawa K, Udagawa H, Mazieres J, et al. Trastuzumab Deruxtecan in HER2-Mutant Non-Small-Cell Lung Cancer. N Engl J Med. 2022;386(3):241–51.

12. Kudek MR, Adashek JJ, Kurzrock R. Ag(e)nostic precision oncology therapy approvals across the years. Trends Cancer. 2025.

13. Marcus L, Lemery SJ, Keegan P, Pazdur R. FDA Approval Summary: Pembrolizumab for the Treatment of Microsatellite Instability-High Solid Tumors. Clin Cancer Res. 2019;25(13):3753–8.

14. Qunaj L, May MS, Neugut AI, Herzberg BO. Prognostic and therapeutic impact of the KRAS G12C mutation in colorectal cancer. Front Oncol. 2023;13:1252516.

15. Wolpin BM, Park W, Garrido-Laguna I, Spira A, Starodub A, Sommerhalder D, et al. Daraxonrasib in Previously Treated Advanced RAS-Mutated Pancreatic Cancer. N Engl J Med. 2026;394(18):1790–802.

16. Meric-Bernstam F. The need for molecular tumor boards. Nature Medicine. 2025;31(10):3228-.

17. Tsimberidou AM, Kahle M, Vo HH, Baysal MA, Johnson A, Meric-Bernstam F. Molecular tumour boards — current and future considerations for precision oncology. Nature Reviews Clinical Oncology. 2023;20(12):843–63.

18. Mosele MF, Westphalen CB, Stenzinger A, Barlesi F, Bayle A, Bieche I, et al. Recommendations for the use of next-generation sequencing (NGS) for patients with advanced cancer in 2024: a report from the ESMO Precision Medicine Working Group. Ann Oncol. 2024;35(7):588–606.

19. Mosele F, Remon J, Mateo J, Westphalen CB, Barlesi F, Lolkema MP, et al. Recommendations for the use of next-generation sequencing (NGS) for patients with metastatic cancers: a report from the ESMO Precision Medicine Working Group. Ann Oncol. 2020;31(11):1491–505.

20. Behel V, Noronha V, Choughule A, Shetty O, Chandrani P, Kapoor A, et al. Impact of Molecular Tumor Board on the Clinical Management of Patients With Cancer. JCO Glob Oncol. 2022;8:e2200030.

21. Scheiter A, Hierl F, Luke F, Keil F, Heudobler D, Einhell S, et al. Critical evaluation of molecular tumour board outcomes following 2 years of clinical practice in a Comprehensive Cancer Centre. Br J Cancer. 2023;128(6):1134–47.

22. Boscolo Bielo L, Guerini Rocco E, Crimini E, Repetto M, Lombardi M, Zanzottera C, et al. Molecular tumor board in patients with metastatic breast cancer. Breast Cancer Res Treat. 2025;210(1):45–55.

23. Schram AM, Fellous M, LeDuke K, Schmid A, Dumbrava EE. PYNNACLE phase II clinical trial protocol: rezatapopt (PC14586) monotherapy in advanced or metastatic solid tumors with a TP53 Y220C mutation. Future Oncol. 2025;21(24):3159–66.

24. Graf T, Boos LA, Mehra T, Miglino N, Sobottka B, Ruschoff JH, et al. Precision Oncology Guided by Genomic Profiling in Breast Cancer: Real-World Data from a Molecular Tumor Board. Cancers (Basel). 2025;17(15).

25. Cortes J, Rugo HS, Cescon DW, Im SA, Yusof MM, Gallardo C, et al. Pembrolizumab plus Chemotherapy in Advanced Triple-Negative Breast Cancer. N Engl J Med. 2022;387(3):217–26.

26. Vandenbroucke JP, von Elm E, Altman DG, Gøtzsche PC, Mulrow CD, Pocock SJ, et al. Strengthening the Reporting of Observational Studies in Epidemiology (STROBE): explanation and elaboration. Epidemiology. 2007;18(6):805–35.

