## Supplemental Figures and Tables for "Effectiveness of a Molecular Tumor Board in Making Personalized Treatment Recommendations for Patients with Advanced Cancer"

Supplemental Figure 1A

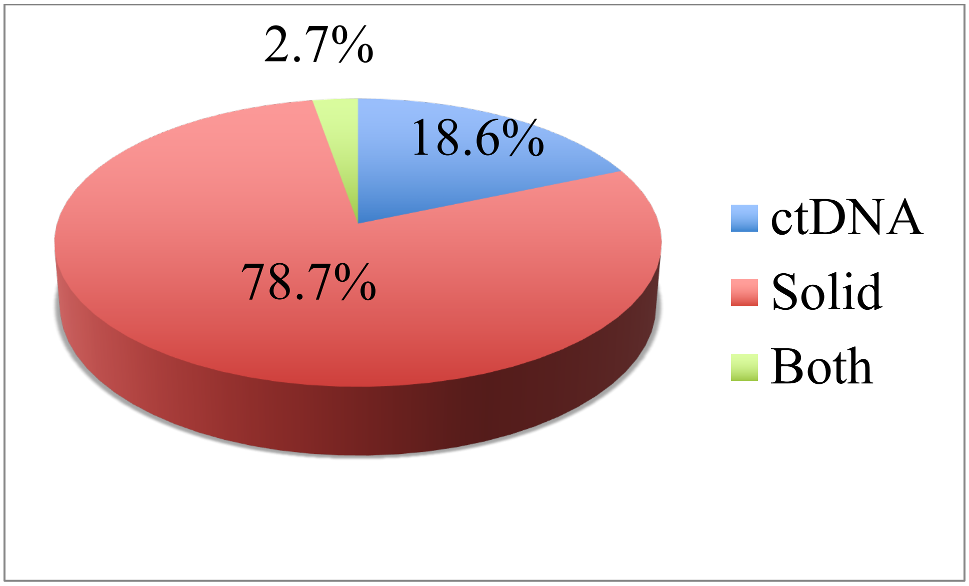


Supplemental Figure 1B

| Patients | N (%)<br>328 (100) |
|---|---|
| **Solid** | 258 (78.7) |
| - Foundation One | 252 |
| - Other | 006 |
| **Liquid** | 061 (18.6) |
| - Foundation One | 057 |
| - Canexia | 002 |
| - Guardant 360 | 002 |
| **Both** | 009 (2.7) |
| - Foundation (TB & LB) | 007 |
| - Foundation (TB) & Canexia (LB) | 002 |

Supplemental Figure 2A

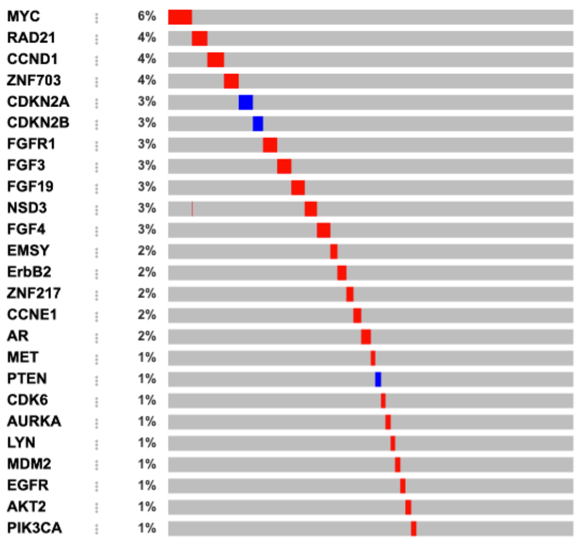


Supplemental Figure 2B

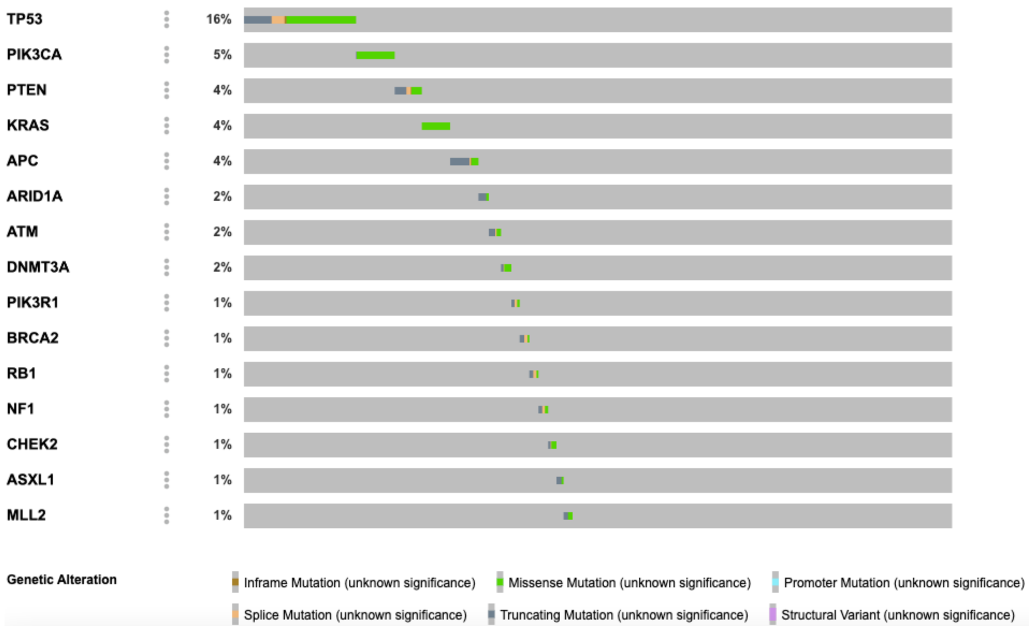

Supplemental Figure 2C

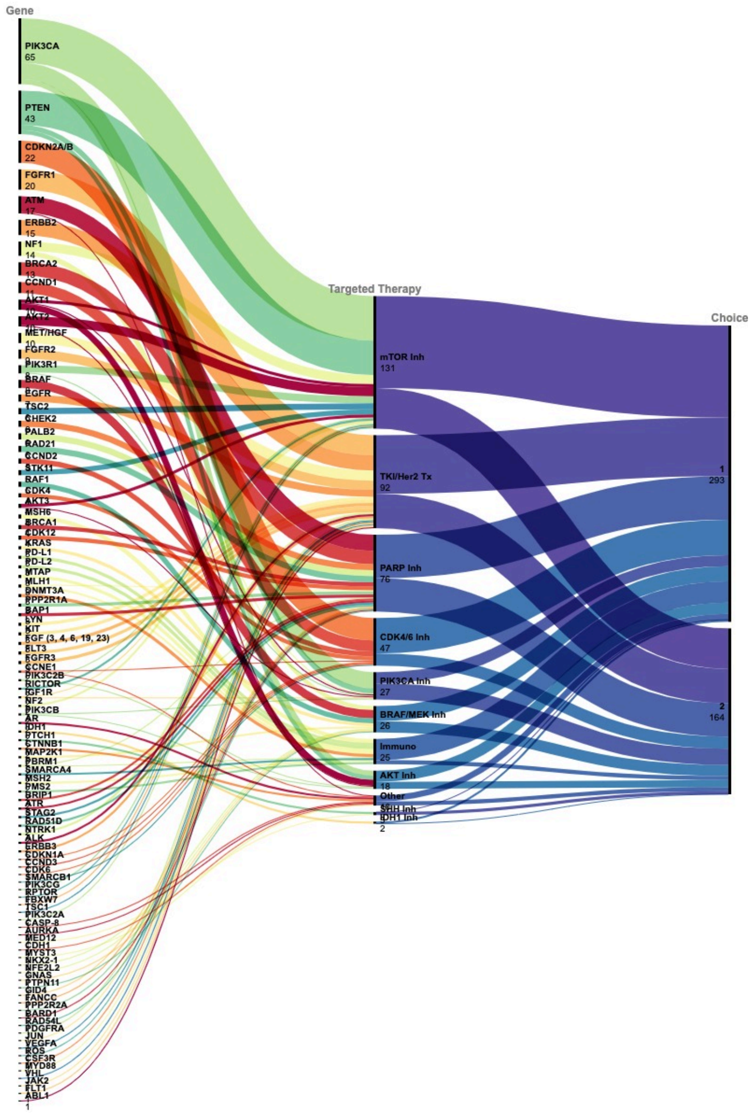

Supplemental Figure 3

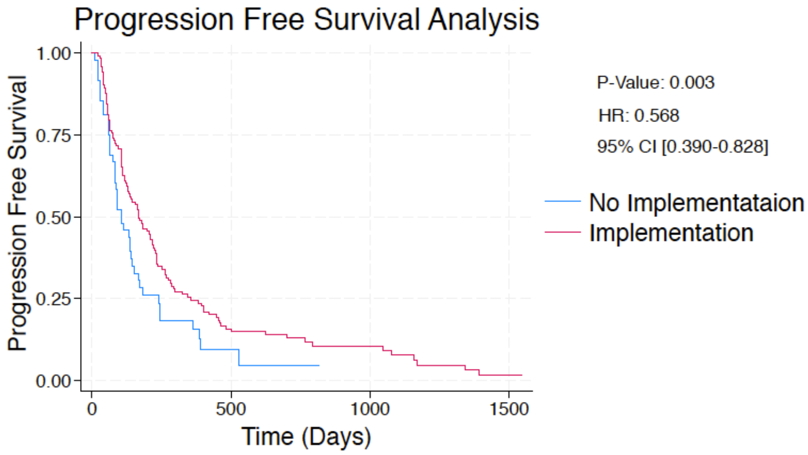

Supplemental Figure 4

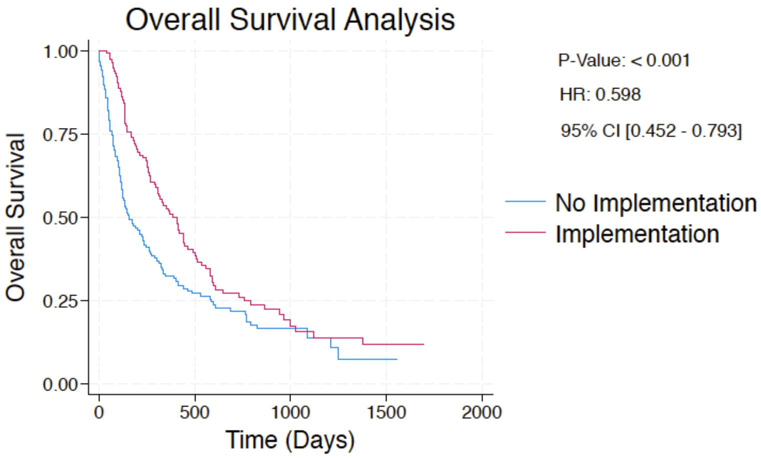

**Supplemental Table 1: 1st MTB Recommended and Implemented Therapies**

| Molecular Alteration | Recommended N (%) | Implemented N (%) |
|---|---|---|
| mTOR Inh | 78 (26.0) | 29 (22.3) |
| PARP Inh | 46 (15.3) | 18 (13.8) |
| TKI | 45 (15.0 | 14 (10.8) |
| Immunotherapy | 37(12.3) | 26 (20.0) |
| CDK4/6 Inh | 30 (10.0) | 18 (13.8) |
| BRAF/MEK Inh | 17 (5.7) | 04 (3.1) |
| AKT/PIK3CA Inh | 17 (5.7) | 08 (6.2) |
| Anti-Her2 Treatment | 15 (5.0) | 10 (7.7) |
| Other | 15(5.0) | 03 (2.3) |
| Total | 300 (100) | 130 (100) |

**Supplemental Table 2: 2nd MTB Recommended and Implemented Therapies**

| Molecular Alteration | Recommended **N (%)** | Implemented **N (%)** |
|---|---|---|
| mTOR Inh | 34 (19.1) | 3 (18.8) |
| PARP Inh | 25 (14.0) | 5 (31.3) |
| TKI | 31 (17.4) | 1 (6.3) |
| Immunotherapy | 15 (8.4) | 3 (18.8) |
| CDK4/6 Inh | 15 (8.4) | 0 (0) |
| BRAF/MEK Inh | 11 (6.2) | 0 (0) |
| AKT/PIK3CA Inh | 17 (9.6) | 2 (12.5) |
| Anti-Nectin ADC | 10 (5.6) | 2 (12.5) |
| Other | 20 (11.2) | 0 (0) |
| Total | 178 (100) | 16 (100) |
